# InpherNet provides attractive monogenic disease gene hypotheses using patient genes indirect neighbors

**DOI:** 10.1101/2020.07.10.20150425

**Authors:** Boyoung Yoo, Johannes Birgmeier, Jonathan A. Bernstein, Gill Bejerano

**Author notes:** **Corresponding Author** Gill Bejerano, Stanford University, Stanford, CA 94305, 1.

## Abstract

Close to 70% of patients suspected to have a Mendelian disease remain undiagnosed after genome sequencing, partly because our current knowledge about disease-causing genes is incomplete. Although hundreds of new diseases-causing genes are discovered every year, the discovery rate has been constant for over a decade. Generating an attractive novel disease gene hypothesis from patient data can be time-consuming as each patient’s genome can contain dozens to hundreds of rare, possibly pathogenic variants. To generate the most plausible hypothesis, many sources of indirect evidence about each candidate variant may be considered. We introduce InpherNet, a network-based machine learning approach to accelerate this process. InpherNet ranks candidate genes based on gene neighbors from 4 graphs, of orthologs, paralogs, functional pathway members, and co-localized interaction partners. As such InpherNet can be used to both prioritize potentially novel disease genes and also help reveal known disease genes where their direct annotation is missing, or partial. InpherNet is applied to over 100 patient cases for whom the causative gene is incorrectly given low priority by two clinical gene ranking methods that rely exclusively on human patient-derived evidence. It correctly ranks the causative gene among its top 5 candidates in 68% of the cases, compared to 9-44% using comparable tools including Phevor, Phive and hiPhive.

## Introduction

Every year, approximately 7 million newborns worldwide are affected by Mendelian diseases^1^. Mendelian diseases are most often monogenic, caused by 1-2 highly penetrant variants in a single gene. In the age of widely available exome sequencing, diagnosing these monogenic conditions can be done by identification of the causal gene against the current body of biomedical knowledge.

one or very few causative genes that contain the disease-causing variants. This is a time-consuming task for clinicians, since exome sequencing can results in hundreds of candidate causative genes that contain variants rarely found in the unaffected population^2–7^. As sequencing technology becomes more time- and cost-efficient, the number of patients being sequenced for disease diagnosis is expected to grow fast. It has been projected that over 60 million patients will be sequenced by 2025^8^.

Numerous tools that automate aspects of the diagnosis pipeline for patients with suspected Mendelian disease have been developed. For example, ANNOVAR^9^ annotates variants with various relevant attributes, and M-CAP^10^ and S-CAP^11^ scores help assess variants’ pathogenicity likelihood. ClinPhen^12^ helps extract patient phenotypes from their free text medical records, and candidate causative gene prioritization tools such as Phevor^13^, PhenIX^14^, Phrank^15^, and AMELIE^16,17^ improve diagnosis efficiency by prioritizing the patient’s candidate for likelihood of causing a patient’s phenotypes. These tools help speed up the diagnosis process, and therefore, allow more cases to be processed.

Despite these technological advances, the clinical diagnostic yield for patients with a suspected Mendelian disease is currently only around 30% after exome sequencing^6^. Hundreds of novel Mendelian disease-causing genes are discovered year in and year out^18–21^. After a novel gene is proved as causative, the relevant gene is conceptually moved from the research realm into the clinic (Figure 1). However, while it is still in the research realm, researchers will consult the biological literature in search of indirect evidence that makes a candidate causative gene a plausible hypothesis for explaining a particular patient’s case. For example, one of the patient candidate genes may have an ortholog known to cause similar phenotypes in a model organism. Similarly, a candidate gene may be in the same functional pathway as known relevant disease-causing genes or have an obligate interaction partner that is already known to cause the patient’s phenotypes.

**Figure 1.**
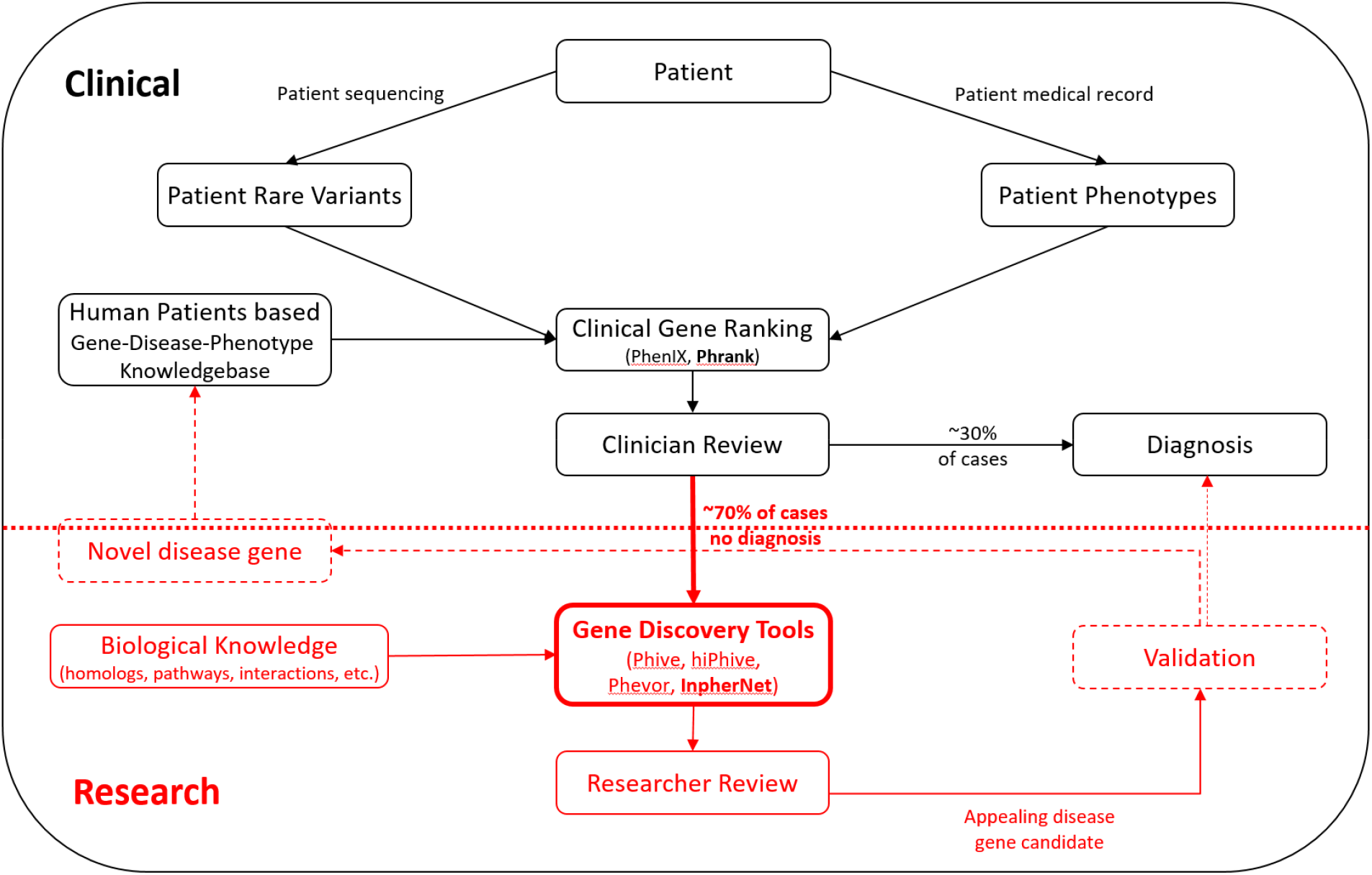
InpherNet’s role in the quest for patient diagnosis. Patient sequencing data is first assessed in a clinical setting, where a diagnosis most often consists of matching a candidate variant or gene in the current patient to previously diagnosed patients with very similar phenotypic abnormalities. When a clinical diagnosis cannot be found (70% of cases), the case moves to the research realm where indirect evidence is sought to suggest a novel causative gene candidate. InpherNet aims to accelerate this discovery process by offering researchers its most appealing testable hypothesis through indirect evidence.

This open-ended search for the most plausible candidate novel disease gene can take months. Since this is an expensive manual task in both time and resources, not all patient cases can be closely scrutinized. Computational inference tools like Phive^14^ and hiPhive^14^ have been developed to help accelerate the discovery of testable research hypotheses. Such tools perform cross-species and gene product interaction-based inferences to prioritize candidate genes beyond patient-based phenotypic knowledge.

Here we propose InpherNet, a network-based machine learning gene prioritization method that automatically sifts through massive amounts of biological information to discover appealing novel disease gene hypotheses. InpherNet leverages indirect evidence not derived from human patients to rank candidate genes without the knowledge of the candidate genes’ functional annotations. InpherNet ignores human patient-derived phenotypes caused by candidate genes to better focus on inferring novel disease genes rather than overweighting those genes with already known phenotype annotations. To predict causative genes using non-patient-derived information, InpherNet considers variant-based information and four sources of indirect evidence, or neighbors: phenotypes associated with orthologs (i.e. the same gene in a different organism), paralogs (i.e. another gene from the same gene family member), members of the same functional pathway, and co-localized interaction partners. Compared to previous diagnosis inference tools, InpherNet constructs an extensive indirect evidence graph, uses a Phrank^15^ based metric to measure set similarity, filters protein interactions by anatomical co-localization, adds variant-related features, and applies a Gradient Boosting Tree classifier to rank the candidate genes for likelihood of causing the patient’s phenotypes based on indirect evidence from candidate genes’ neighbors. We test our performance on 137 previously diagnosed patients whose causative genes were incorrectly given low rankings by clinical tools that rely on patient-derived phenotypic data. In such cases, InpherNet outperforms three existing inference tool configurations and three clinical tool configurations in ranking causative genes based on indirect evidence, thus showing its potential to accelerate research efforts targeted towards the discovery of novel testable disease-causing genes and ultimately increasing diagnostic yield.

## Materials and Methods

### InpherNet Graph

#### Mendelian subgraph of Monarch Initiative’s multi-species biological network

The Monarch Initiative^22^ is an effort to develop a comprehensive biological database containing numerous entities (e.g. genes and phenotypes) and relationships between these entities from multiple existing databases (e.g. gene functions from Gene Ontology^23^ and disease-causing genes from Orphanet^24^). Since InpherNet aims to prioritize candidate genes in patients affected with Mendelian diseases, we defined a subset of the Monarch graph database that is both relevant to Mendelian disease inference and annotated with sufficient data (see Supplementary Methods). The Monarch Initiative graph knowledgebase is particularly appealing because it is built using unified ontologies such as Uberon^22^ and Upheno^22^ that greatly facilitate the comparison of cross-species observations. It also includes Human Phenotype Ontology associations^25^ (HPO-A) which contain gene-phenotype causative relationships. Phenotypes are organized in a directed acycylic graph of phenotypic abnormalities, the Human Phenotype Ontology (HPO). Gene-phenotype causative relationships are curated from the database Online Mendelian Inheritance in Man (OMIM), a database containing manually curated facts about Mendelian diseases and their causative genes, and similar databases. In total, we selected 9 ontology sources (see Supplementary Table 1) encompassing 1,231,846 attributes of human, mouse, and zebrafish genes (see Figure 2).

**Table 1.**
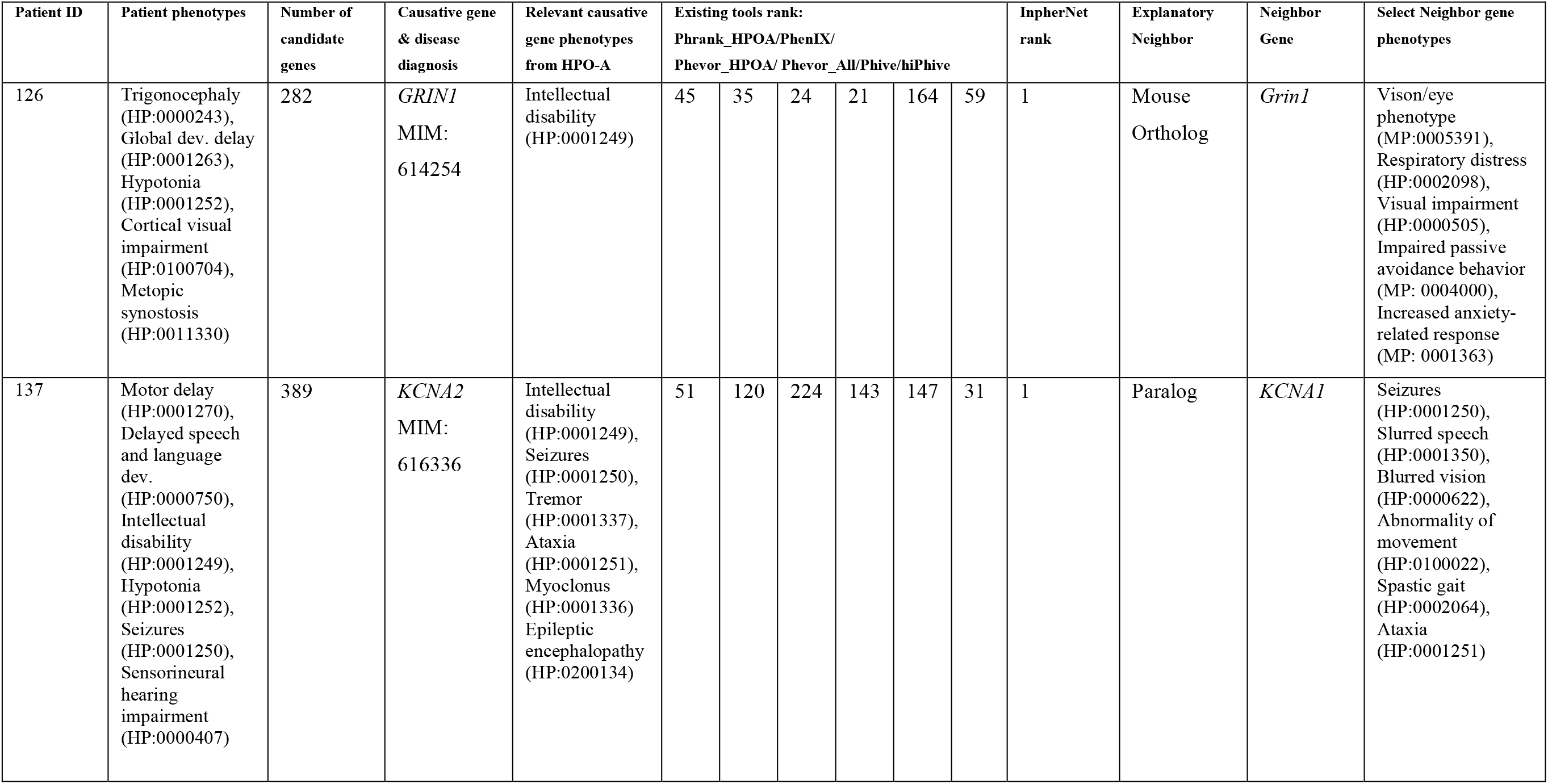
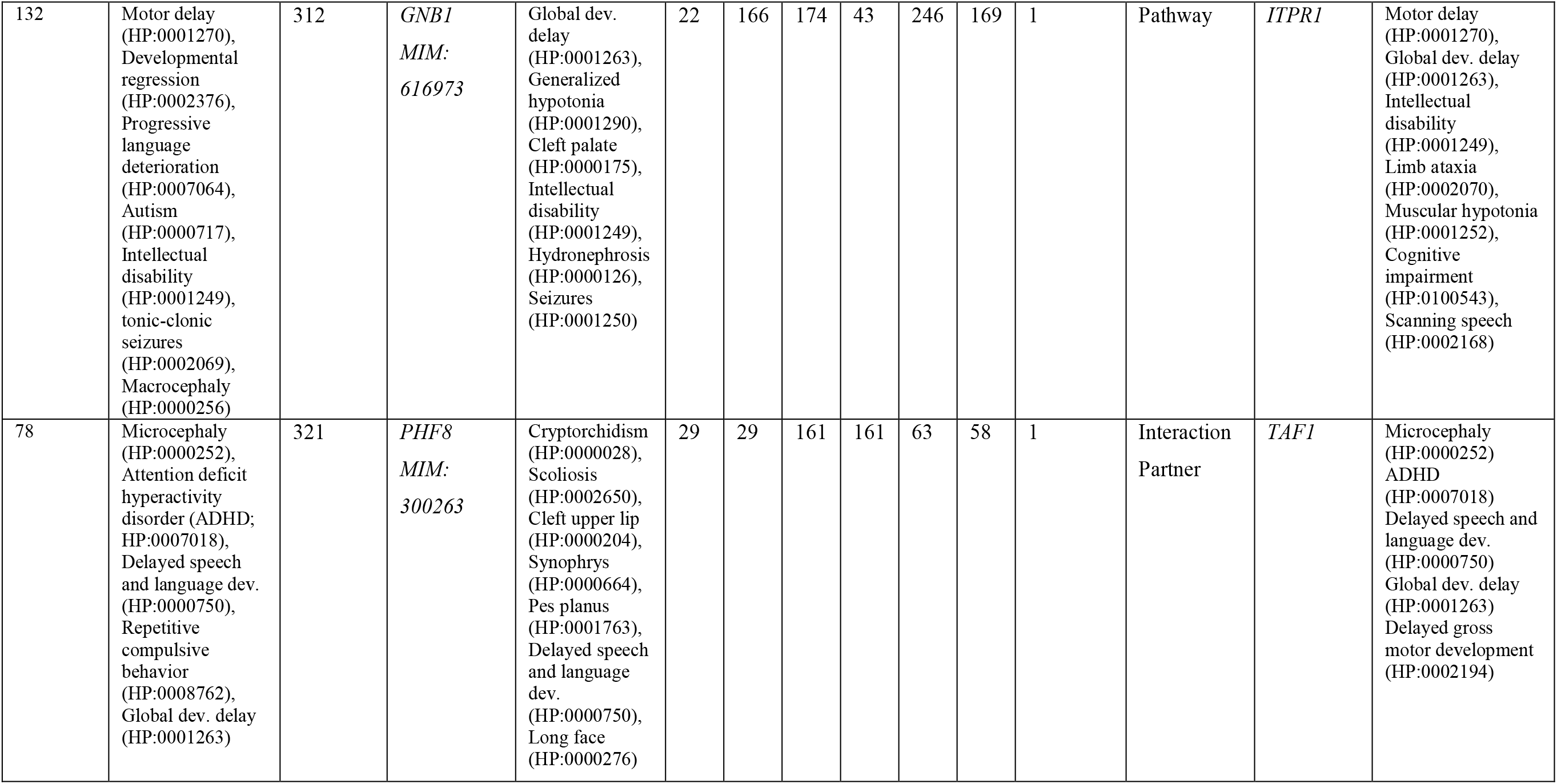
InpherNet supports each prediction with referenced observations. To allow researchers to easily evaluate its predictions, InpherNet outputs supporting information for each candidate gene. Here, we show examples where InpherNet outperformed all other tools, along with supporting phenotypic evidence and its source for its highest Phranken scoring explanatory neighbor.

**Figure 2.**
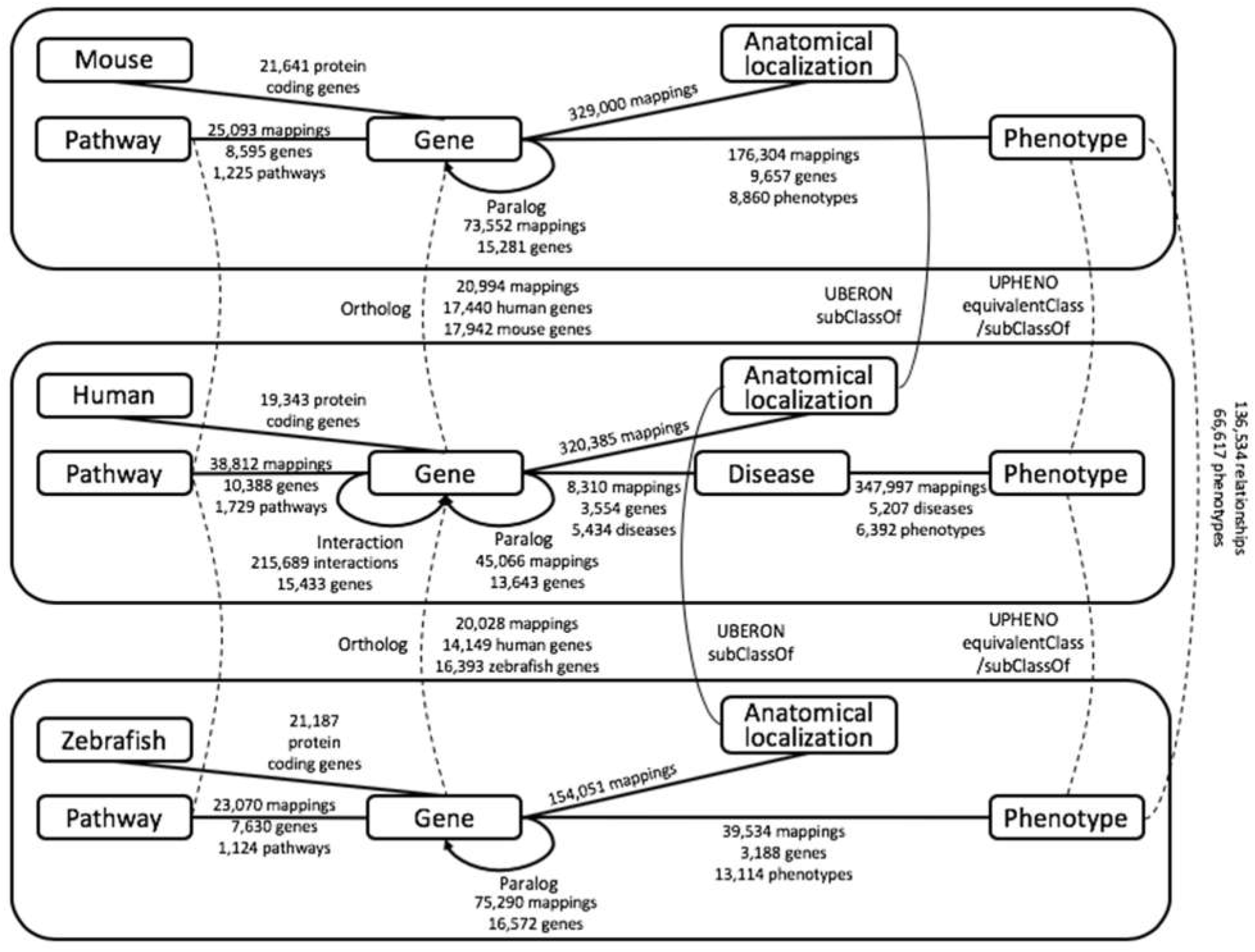
The multi-modal biological network underlying InpherNet. We extract human-, mouse-, and zebrafish-based orthology, paralogy, pathway, interaction, anatomical localization, phenotypes and monogenic disease relationships from Monarch Initiative’s graph database and Ensembl.

#### Gene orthology and paralogy mappings from Ensembl

Ensembl^26^ is a consortium that develops and curates many resources for comparative genomic analyses. We used its human, mouse, and zebrafish gene orthology and paralogy relationships in the InpherNet graph (see Figure 2). Extending phenotypic abnormalities associated with human genes through their mouse and zebrafish orthologs enabled hypothesis generations on many more human genes than is currently possible with just human data. For example, only 3,438, or 17.8%, of human protein-coding genes in our graph are annotated with direct human patient-derived phenotypes. However, after projecting mouse and zebrafish phenotypes to their orthologous human genes, over 56.2% (3.2x more) of human genes can be phenotypically annotated (see Figure 3). In our dataset, 17,784 genes out of 19,343 total human genes (91.9%) have orthologous genes in either or both mouse or zebrafish, 13,315 (68.8%) have in-paralogous genes, and 13,189 (68.2%) have out-paralogous genes in either or both mouse or zebrafish. If we consider phenotypic information projected from orthology, in-paralogy, and out-paralogy in human, mouse, and zebrafish, the number of human genes that can by phenotypically annotated increase to 71.8% (4.04x more).

**Figure 3.**
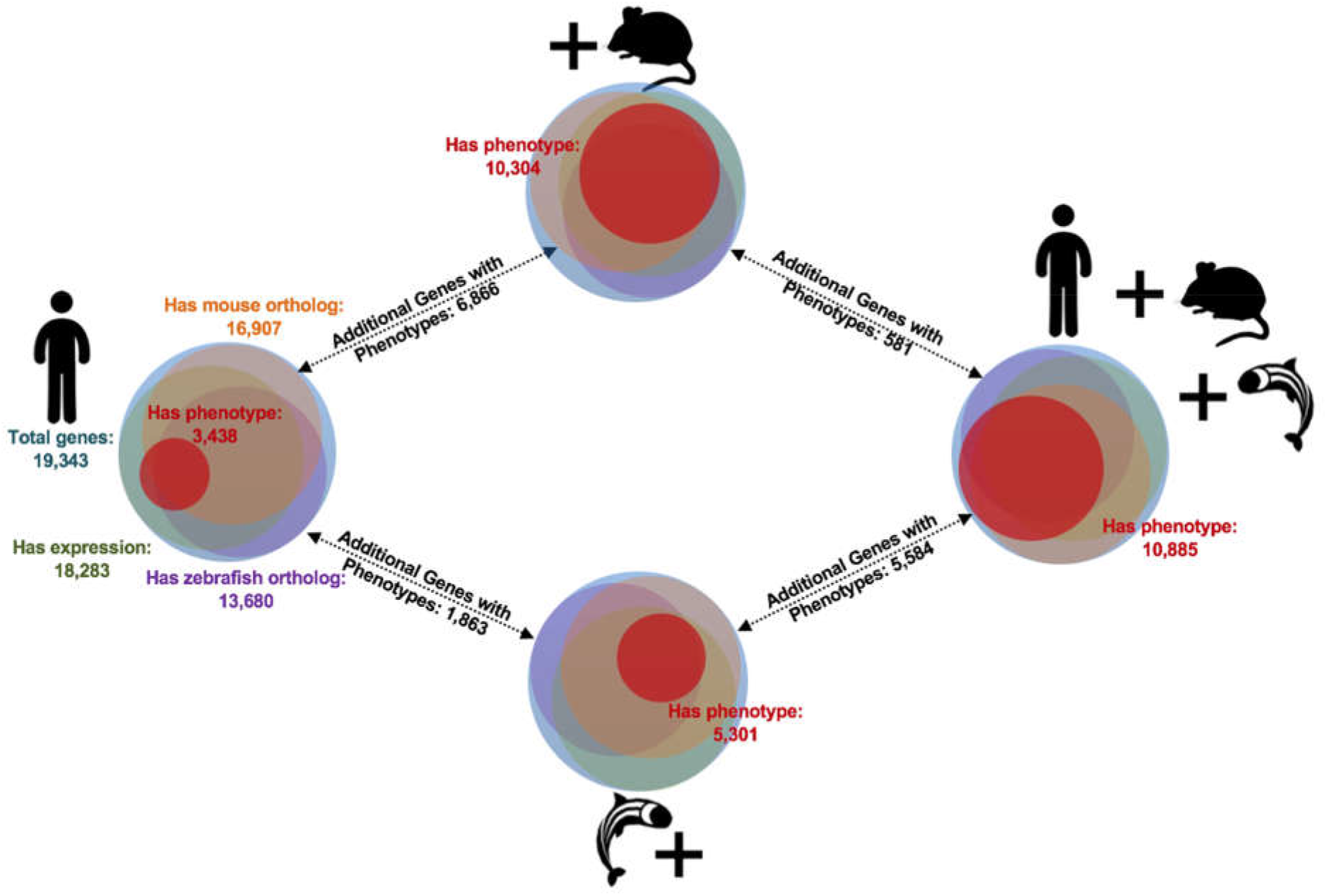
The power of orthology. HPO-A contains human gene-phenotype relationships for about 3,400 out of 20,000 human protein-coding genes. Thus, clinical gene prioritization methods that only use known human Mendelian gene-phenotype associations cannot prioritize 82.4% of human protein-coding genes. However, many unannotated human genes have functionally annotated orthologs in mouse and zebrafish that can be combined via Monarch Initiative’s Upheno cross-species phenotype ontology to triple the annotation coverage to 56% of human protein-coding genes compared to the original 17.5%.

### Gene scoring by means of a supervised machine learning algorithm

Our machine learning classifier takes a vector of scalar values (called “features”) as input, and outputs a score between 0 and 1, indicating the classifier’s assessment of whether the input should be classified as positive (here, indicating that the indirect evidence suggests a gene is causative for a patient) or negative (here, that the indirect evidence does not support a match). InpherNet used a Gradient Boosting Tree^27^ classifier, a type of supervised machine learning classifier, to assign such a score to each candidate gene (see Figure 4 and Supplementary Methods).

**Figure 4.**
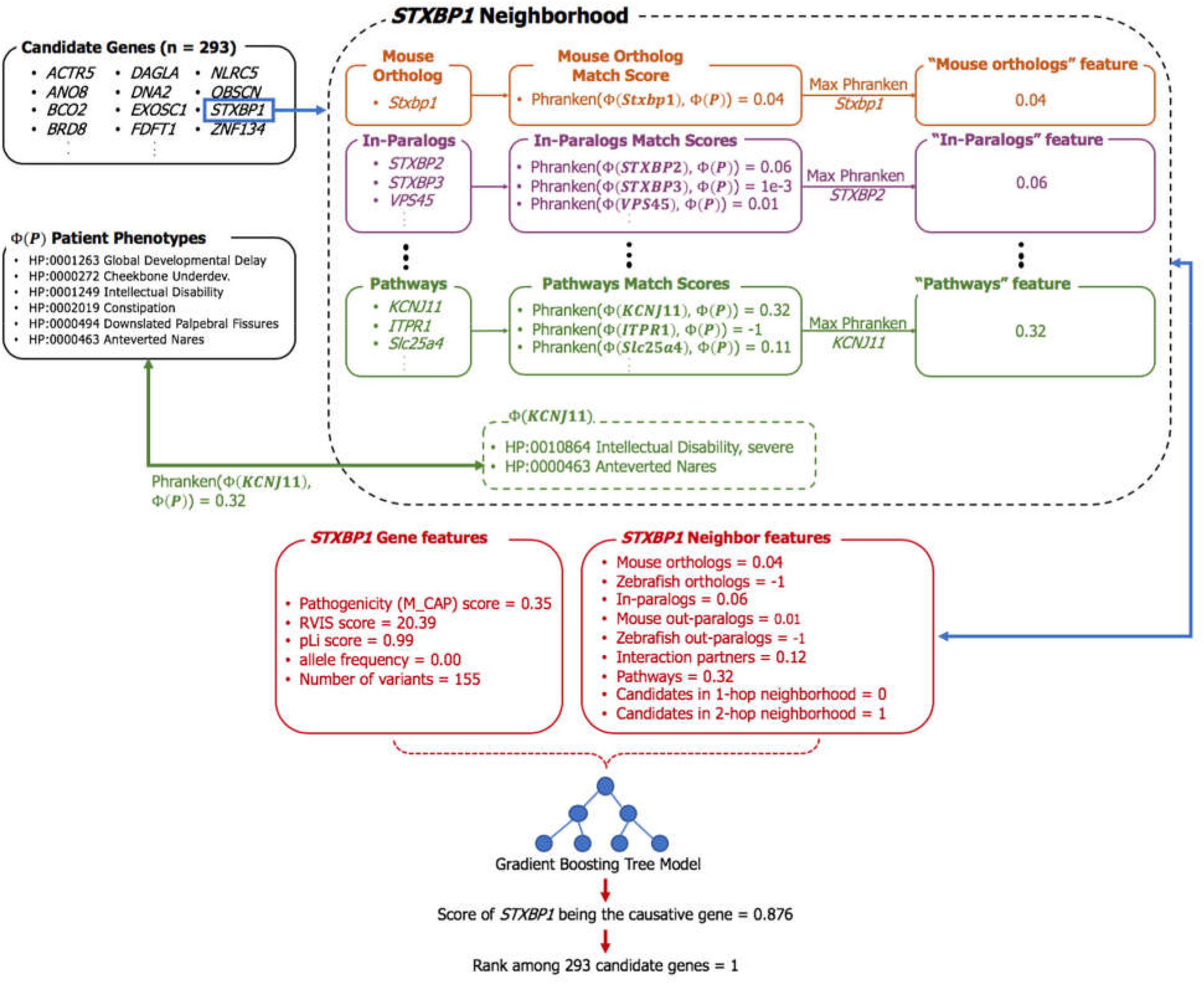
InpherNet’s graph-based machine learning classifier. InpherNet aggregates a patient’s candidate genetic variants to a list of candidate genes. These Phranken (Phrank-normalized) scores along with additional gene and variant level information are passed into a Gradient Boosting Tree classifier, which returns a score between 0 and 1 quantifying the candidate gene’s likelihood of being causative. InpherNet then ranks all candidate genes on this score. For each gene, we find its set of neighbors in one of the seven contexts, and pick the highest Phranken match score between a neighbor’s phenotypes and the patient’s phenotypes per context (or −1 when no neighbor has known associated phenotypes).

### InpherNet feature set

Candidate genes are genes that contain at least one candidate causative variant in the patient (see Supplementary Methods). We associated each candidate gene with a vector of 15 features derived from information about the candidate gene’s neighbors (orthologs, paralogs, pathways, and interaction partners) and the candidate variants. (see Figure 4). None of the 15 features are about the direct functional annotations of the candidate gene, which prevents overweighting candidate genes with previously known patient-derived phenotypes, since InpherNet aims to derive hypotheses for novel disease-causing genes from indirect evidence.

### Gene neighbors feature set

#### Mouse ortholog

Orthologs are similar genes in two different species related via a speciation event, and often have similar functions^28^. For each patient candidate gene, we computed a phenotypic match score between the mouse ortholog-associated phenotypes from the Mouse Genome Informatics (MGI) phenotype database^29^ and the patient phenotypes in HPO terms using a Phrank^15^-based phenotype match score that we call “Phranken” (for Phrank-normalized). The Phranken score takes two sets of phenotype terms and an underlying phenotype directed acyclic graph (DAG) as inputs and quantifies how similar the two sets are with a single match score (see Supplementary Methods). In InpherNet, we took the phenotype DAG and gene’s phenotype annotations from Upheno^22^, a Monarch Initiative unified phenotype ontology that merges terms from multiple sources including HPO-A, Gene Ontology, MGI Phenotype, and Zebrafish Information Network (ZFIN). If a candidate gene has more than one mouse ortholog, we computed the Phranken match score for all mouse orthologs and selected the ortholog that has the highest Phranken score as the value for the “mouse ortholog” feature. If a candidate gene has no mouse ortholog or none of the mouse orthologs has any known function, −1 is assigned. The same convention is used repeatedly to compute the other neighbor feature values described below.

#### Zebrafish ortholog

Defined as the highest Phranken match score between zebrafish ortholog-associated phenotypes from ZFIN and the patient’s phenotypes.

#### Human in-paralog

In-paralogs are genes found in the same species that are in the same gene family. For human patient-derived phenotypes in HPO-A, their phenotype abnormalities are linked through a disease term from OMIM^30^ or Orphanet^24^ (see Figure 4). Therefore, for human genes, instead of calculating the max Phranken score per gene, we calculated the max score per disease (see Supplementary Methods). For each candidate gene, we computed the Phranken scores for all diseases known to be caused by all human in-paralogs and picked the highest score.

#### Mouse out-paralog

For each candidate gene, we collected all mouse in-paralogs of the candidate gene’s mouse ortholog, which are also known as mouse out-paralogs. The highest Phranken match score between mouse out-paralog-associated phenotypes and the patient’s phenotypes is picked.

#### Zebrafish out-paralog

We similarly used the candidate gene’s zebrafish out-paralogs.

#### Pathway

For each patient candidate gene, we collected all human, mouse, and zebrafish genes that are in the same Reactome^31^ pathways as the patient candidate gene. For human genes, we also collected diseases they are known to cause and their related phenotypes. The highest Phranken match score between the patient’s phenotypes and any pathway gene’s phenotypes for mouse and zebrafish genes or pathway gene’s disease phenotypes for human genes is then used.

#### Interaction partner

For each candidate gene, we retrieved a set of interaction partners supported by both a human protein-protein interaction (PPI) BioGRID^32^ network and human anatomical localization Uberon^22^ data (see Supplementary Methods). Intuitively, we searched for genes whose protein products may interact with the candidate genes in human cells. We picked the highest Phranken score between the patient’s phenotypes and the phenotypes related to the diseases the interaction partners are known to cause.

#### Candidates in 1-hop neighborhood

For each candidate gene, we defined a 1-hop neighborhood as a set of genes that can be reached through 1-hop interaction links from the human PPI network defined above (i.e. their gene products can interact directly in human cells). We counted how many other candidate genes are in this neighborhood, and this count is reported as the “candidates in 1-hop neighborhood” feature.

#### Candidates in 2-hop neighborhood

For the “candidates in 2-hop neighborhood” feature, we repeated the step above but looked for 2-hop neighborhood instead. The 2-hop neighborhood excludes all genes in the 1-hop neighborhood.

## Variant-based feature set

### M-CAP^gene^

M-CAP^10^ is a pathogenicity score that assigns a number between 0 (likely benign) and 1 (possibly pathogenic) to rare human missense variants. We calculated an M-CAP-based feature for each gene as the maximum M-CAP score of all candidate causative variants in the candidate gene. A candidate variant that did not have an M-CAP score were assigned the maximum M-CAP score in a window of −50, +50 basepairs adjacent to that variant.

### M-CAP^100^

We calculated the highest M-CAP score in a window of −50, +50 basepairs adjacent to all candidate variants in the candidate gene, then used the maximum of those values as the value for this feature.

### RVIS score

This is the RVIS^33^ gene mutability score of the candidate gene.

### pLI score

This is the pLI^34^ haploinsufficiency score of the candidate gene.

### Average ExAC allele count

The average Exome Aggregation Consortium^34^ (ExAC) allele count of all candidate variants in the candidate gene is used.

### Candidate variants count

We set this to the number of variants in the candidate gene. For M-CAP, RVIS, pLI, and ExAC, if the original resource did not offer relevant values, we assigned a default “null” value (see Supplementary Methods).

### Other gene prioritization tools

We compared InpherNet’s performance to 6 existing gene prioritization tool configurations. Phrank_HPOA^15^ and PhenIX^14^ are designed for clinical use and target genes that have patient-based Mendelian disease associations, while Phevor^13^, Phive^14^ and hiPhive^14^, similar to InpherNet, use non-patient-based functional information and serve as inference tools for novel disease-gene candidates. Phrank_HPOA ranks candidate genes by their Phrank match score using gene annotations from the HPO-A database. PhenIX ranks candidate genes by their human phenotypic annotations semantic similarities with the patient’s phenotypes in HPO and the candidate causative variants’ pathogenicity. Phevor^13^ combines phenotype, disease, and functional ontology information to rank patient candidate genes. For the comparison to InpherNet, we used Phevor in two ways: “Phevor_HPOA” uses only patient-based phenotypic annotation (HPO-A), similar to the clinical tools, and “Phevor_all” uses all available ontologies similar to the other inference tools. Phive ranks candidate genes using mouse phenotypic data, and most comparable to InpherNet, hiPhive combines functional data derived from human, mouse, and zebrafish genes and the candidate genes’ relatedness in a PPI network to the suspected causative gene to rank candidate genes. We took great care to compare the causative gene ranking performance of all methods on equal footing (see Supplementary Methods for more details).

### InpherNet training set

In order to conserve real patient data for testing (below), we constructed a set of synthetic patients to train the InpherNet’s Gradient Boosting Tree classifier. For this process, we used 2,504 sequenced individuals who are not affected by Mendelian diseases from 1000 Genomes Project (KGP)^35^, and Mendelian disease-causing variants from ClinVar^36^. ClinVar associates pathogenic variants with a disease identifier from OMIM. Further, HPO-A contains HPO phenotypes associated with an OMIM disease. To construct a synthetic patient, we took a KGP genome, added one randomly selected disease-causing genetic variant from ClinVar for a known OMIM disease, and associated the patient with a set of noisily sampled disease-associated phenotypes, mimicking imperfect clinical annotations (see Supplementary Methods). We also ensured that no causative gene in the training set was equal to the causative gene of any real patient used for testing or validating to prevent overfitting. Using this method, we generated 2,504 different synthetic patients with an average of 9.1 phenotypes and 300.4 candidate genes per patient.

### InpherNet test set

We tested InpherNet’s performance on 137 real patients with their pre-diagnosis phenotypes and clinician-verified Mendelian diagnoses (see Supplementary Methods). Since InpherNet is a novel gene function inference tool rather than a clinical tool, we created a test set containing diagnosed patient cases whose causative genes were incorrectly given low priority by clinical gene-ranking tool. This was done to illustrate InpherNet’s performance on cases where a clinician was not able to find a diagnosis by examining the genes ranked highest by clinical tools, and then consulted an inference tool in search for novel diagnosis hypothesis (Figure 1). We first ran PhenIX and Phrank_HPOA on all available 255 patients to find a cohort of patients where PhenIX failed to rank the causative gene among its top 10 output genes (*PhenIX > 10*), and a cohort of patients where Phrank_HPOA failed to rank the causative gene among its top 10 output genes (*Phrank > 10*; see Supplementary Table 2). The test cohort *PhenIX > 10* contains 115 patients (with an average of 7.8 phenotypes and 309.0 candidate genes per patient) and the test cohort *Phrank > 10* contains 88 patients (with an average of 8.0 phenotypes and 314.4 candidate genes per patient). Since in the real clinical diagnosis process, the top 10 genes from these clinical tools would already have been scrutinized by a clinician and discarded as non-causative, we removed the top 10 PhenIX- or Phrank_HPOA-ranked genes from the candidate gene list for each patient in both test cohorts *PhenIX > 10* and *Phrank > 10*, respectively.

### Interpreting InpherNet gene rankings

To provide human-interpretable explanations for InpherNet’s gene rankings, we included a ranked list of each candidate gene’s neighbors ordered by the Phranken phenotype match score between the patient’s phenotypes and neighbor-associated phenotypes. This list helps researchers see which neighbor is phenotypically most similar to the patient’s phenotypes, and via the Monarch Initiative subgraph we used in InpherNet, link back to the original databases supporting these claims.

## Results

### InpherNet outperforms existing phenotype ranking-based methods

The goal of candidate gene ranking tools is to rank the true causative gene at the top to allow clinicians find diagnoses, or enable researchers to propose a novel hypothesis, after reviewing as few candidate genes as possible. As such, we used the causative gene’s rank among all patient candidate genes to evaluate the performance of gene rankings methods (see Figure 5).

**Figure 5.**
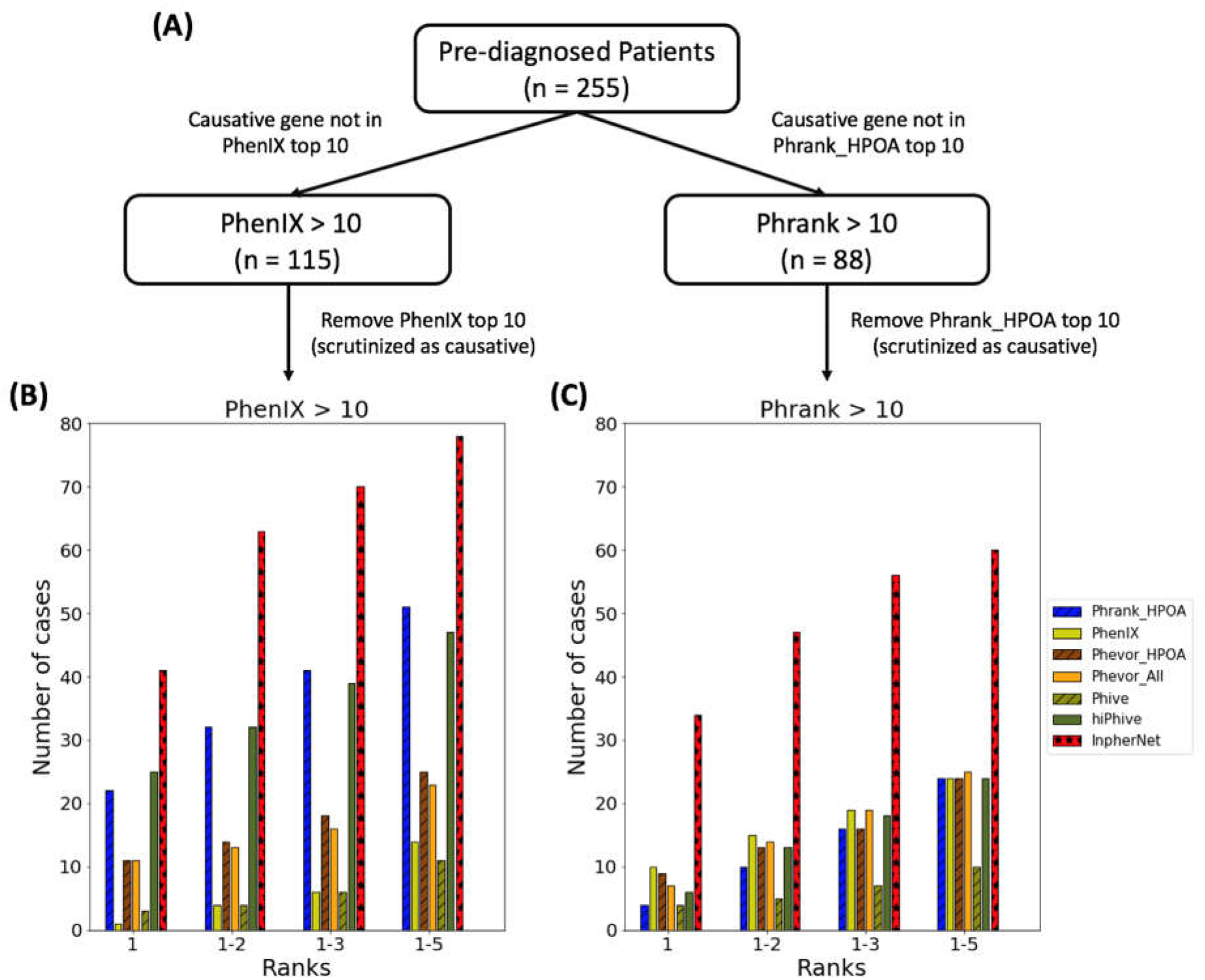
InpherNet outperforms existing gene prioritization methods. To mimic the clinical transition to the research inference realm (Figure 1), we took 255 patients with diverse pre-diagnosed conditions and first used two human data-only based methods to attempt quick clinical diagnosis. For 137 out of 255 patients, the causative gene was not among the top 10 output genes by at least one of the two tools. **(A, left)** In 115 (83.9%) of 137 cases the causative genes was not among the top 10 PhenIX-prioritized candidate genes. **(A, right)** In fewer cases (88 of 137, 64.2%), the causative gene was not in the Phrank_HPOA top-ranked 10 genes. Each case is then reviewed by multiple inference tools after removing the highest ranked 10 genes that were already determined to be not causai. **(B)** 7 different gene prioritization tools configurations were run on the remaining candidate genes (3 clinical and 4 inference tools). Since the top 10 PhenIX genes have been removed, PhenIX can still rank the causative gene high among the remaining candidate genes. However, the graph shows that most of the cases are truly difficult for the clinical tools, and the causative gene is not getting ranked just beyond the top 10 limit. **(C)** The same measurements were taken following the clinical use of Phrank_HPOA instead of PhenIX. In both scenarios InpherNet outperforms all previous methods, ranking more causative genes higher based on neighbor information alone.

In *PhenIX > 10*, the set of 115 real test patients where PhenIX ranks the causative gene worse than 10, InpherNet ranks the causative gene among the top 5 output candidate genes for 68.7% of patient cases, while the 3 existing inference tools we compared to prioritize the causative gene in the top 5 output candidate genes in 40.9% (hiPhive), 20.0% (Phevor_All), 9.6% (Phive) cases, and the 3 clinical tools do so in 44.3% (Phrank_HPOA), 12.2% (PhenIX), and 21.7% (Phevor_HPOA) cases. InpherNet outperforms all other methods even further in *Phrank > 10*, the set of 88 real test patients whose causative gene has a Phrank_HPOA rank worse than 10, where InpherNet ranks the causative gene among the top 5 candidates in 68.2% of the cases compared to 27.3% (hiPhive), 28.4% (Phevor_All), 11.4% (Phive) by the existing inference tools and 27.3% (Phrank_HPOA), 27.3% (PhenIX), 27.3% (Phevor_HPOA).

### InpherNet ranks candidate genes that lack patient phenotype annotations

Ranking candidate genes that lack any patient phenotype annotations is critical for the discovery of novel disease-causing genes. In our real patient test cohort *Phrank > 10*, an average of 77.3% of patient candidate genes (8,204 unique genes across all 88 patients) do not have any HPO annotations and are therefore automatically ranked at the bottom by methods that rely only on HPO-A (see Supplementary Methods). In two real diagnosed patient cases out of 88 in our *Phrank > 10* test set, their causative genes do not have any HPO annotations. InpherNet is able to rank the causative gene relatively high for both cases. Phrank_HPOA ranks that causative gene *BPTF* top 190 among 311 candidate genes along with all other candidate genes lacking HPO annotations for Patient:131, but InpherNet ranks BPTF top 10. For Patient:092, Phrank_HPOA assigns the causative gene *CHD8* rank 198 out of 346 candidate genes while InpherNet improves this rank to top 36.

### Interpretability of InpherNet’s prediction process

All four neighbor types (orthology, paralogy, pathways, and interactions) contributed to InpherNet’s performance. Table 1 provides an example each where InpherNet used phenotypic evidence from each type of candidate gene neighbors to rank the causative gene higher than other gene prioritization methods.

For example, in Patient:126, the causative gene *GRIN1* is ranked at the top by InpherNet through its mouse ortholog *Grin1*. In HPO-A *GRIN1* is annotated by only one broad phenotype term, Intellectual disability (HP:0001249), resulting in a low Phrank_HPOA rank of 45 among 282 candidate causative genes. However, InpherNet ranks *GRIN1* at the very top because its mouse orthologous gene, *Grin1*, is well annotated with relevant phenotype such as Visual impairment (HP:0000505).

Similarly, InpherNet ranks the causative gene *KCNA2* for Patient:137 at the top of 389 candidate genes through information about its in-paralog, *KCNA1*. Human patients with rare variants in *KCNA1* have shown phenotypes similar to this patient’s phenotypes including Seizures (HP:0001250), Slurred speech (HP:0001350) and Abnormality of movement (HP:0100022). The patient’s actual causative gene, *KCNA2*, lacks motor- or speech-related patient-based phenotype annotations resulting in low rank (51 for Phrank_HPOA and 120 for PhenIX) for the clinical ranking tools, and inference tool hiPhive ranks this gene in top 31.

In the case of Patient:132, the causative gene identified is *GNB1*. Phrank_HPOA ranks this gene at 22, and hiPhive 169. InpherNet ranks this gene at the top among 312 candidate causative genes predominantly through *ITPR* which is in the same Ca^2+^ pathway^31^ as *GNB1*.

Finally, the connection between interaction partners *PHF8* and *TAF1* bring the correct causative gene *PHF8* for Patient:078 to the top, while Phrank_HPOA ranks *PHF8* at position 29 and hiPhive at position 58 among 321 candidate genes. While rare mutations in both *PHF8* and *TAF1* are known to cause X-linked mental retardation in patients, the phenotypic abnormalities associated with each of these genes in our knowledgebase differ. The knowledgebase associates *PHF8* with phenotypic abnormalities unobserved in this patient including Long face (HP:0000276) and Cleft upper lip (HP:0000204). However, its well-known interaction partner *TAF1* is associated with much more relevant phenotypes including Microcephaly (HP:0000252) and Attention-deficit/hyperactivity disorder (HP:0007018).

## Discussion

Extensive efforts are being made to diagnose the growing number of sequenced patients with suspected Mendelian diseases^2,3,13–16^. However, 70% of sequenced patients with suspected Mendelian diseases remain undiagnosed^6^, partly due to incomplete knowledge of Mendelian disease-causing genes. While novel disease-causing genes are continuously being identified, the discovery rate has been steady at around 250 genes per year for over a decade^18–20^. Here we show that InpherNet improves the ability to navigate underexplored areas in the gene-phenotype space by harnessing existing knowledge of four biologically significant neighbor relationships—orthology, paralogy, pathway membership, and interaction partners—to help accelerate the discovery of novel disease-causing genes. These neighbors provide additional evidence that is not yet directly linked with candidate genes, thus suggesting appealing testable candidates.

InpherNet currently uses mouse and zebrafish orthologs and their phenotypic annotations to expand known human genes’ patient-based phenotype annotations. We show that from these two species, we can increase the number of phenotypically annotated human genes four-fold. While we wish to include additional species in InpherNet, the next best phenotypically annotated species in the Monarch Initiative data, rat, only has 1,231 gene-phenotype relationships compared to zebrafish, 42,367 relationships, and mouse, 184,313 relationships. As additional species’ functional data becomes more complete, they can be easily integrated into InpherNet’s flexible Gradient Boosting Tree model.

InpherNet accelerates the discovery process of monogenic disease-causing genes by integrating information about neighboring genes that are associated with patient-similar phenotypes. In contrast, clinical gene prioritization tools such as Phrank_HPOA^15^ and PhenIX^14^ rely exclusively on human-observed phenotypes. While Phevor^13^, Phive^14^, and hiPhive^14^ also consider non-human functional annotations, we show that InpherNet improves performance specifically on cases where clinical tools do not bring the causative gene near the top. Although our performance was measured on previously diagnosed patients, we show that InpherNet also ranks the causative gene high for cases whose causative genes are lacking any patient-derived phenotype annotations. In addition, InpherNet is able to effectively rank the causative gene without any knowledge of candidate genes causative diseases which shows it is able to discover disease-causing genes. Our results also corroborate the finding from the Phrank^15^ paper on its ability to improve on PhenIX, as in more of our cases the causative gene is ranked worse than top 10 by PhenIX than by Phrank_HPOA. Furthermore, Phrank_HPOA ranks the causative genes in top 5 for most cases excluding InpherNet in *PhenIX > 10*, while PhenIX does not perform as well in *Phrank > 10*.

By integrating indirect biological knowledge to infer novel gene functions and to provide novel hypothesis for further validation, InpherNet improves the state of the art to advance the forefront of clinical knowledge for Mendelian genetics.

## Data Availability

A portion of the data we use is available from EGA. Another portion is of consented Stanford patients. Some of the latter can be shared while respecting consent conditions.

## Declaration of Interests

The authors declare no competing interests.

### Acknowledgements

We thank members of the Bejerano laboratory, particularly Karthik Jagadeesh, Cole Deisseroth, Ethan Dyer, Heidi Chen, Ethan Steinberg, Surag Nair, Yosuke Tanigawa, Rachel Mangles and Helen Kim for technical advice and valuable feedback; Monarch Initiative members Melissa Haendel, Christopher Mungall, Jeremy Xuan and Kent Shefchek for providing access to and guidance in using Monarch Initiative data; Maria Haanpaa, as well as Jennefer Kohler, Devon Bonner and their colleagues for advice.

## Web Resources

InpherNet code will be available upon publication at https://bitbucket.org/bejerano/inphernet/

